# Extending Problem and Solution Trees: A Participatory Method for Community Partnership Development

**DOI:** 10.64898/2026.10.29.26366845

**Authors:** Nadia A. Sesay, Kendra OoNorasak, Mansura Shahad Bawa, Deidra Haskins, Kristin Hughes, Brandi M. White

**Affiliations:** College of Health Sciences and College of Communication and Information, University of Kentucky, Lexington, Kentucky, United States; College of Health Sciences, University of Kentucky, Lexington, Kentucky, United States; Martin-Gatton College of Agriculture, Food and Environment, University of Kentucky, Lexington, Kentucky, United States; FoodChain, Lexington, Kentucky, United States

**Author notes:** **Corresponding Author**, Brandi M. White, PhD, MPH, 900 South Limestone Street, Lexington, Kentucky 40536-0200.

## Abstract

**Background:** Problem and solution trees (PASTs) are a participatory method for understanding complex environmental and systemic health problems and incorporating community input in appropriate solutions. While PASTs have been applied in international contexts to examine critical health concerns (e.g., hand-washing behaviors and malnutrition), their use in the United States (US) is limited.

**Objectives:** Together with a Project Advisory Board (PAB), identify barriers to DASH (Dietary Approaches to Stop Hypertension) eating plan adoption and co-develop actionable solutions to low adherence for a historically marginalized community in the U.S.

**Methods:** PASTs are visual tools. Over virtual and in-person meetings, we constructed and refined five distinct trees to address each core barrier to DASH adherence. We made real-time edits to trees in PowerPoint and engaged in group discussion to reach consensus on barriers and solutions. Supporting material included a project workbook distributed to the PAB. Meetings were audio-recorded, and transcripts were reviewed following each meeting to ensure the accuracy of each tree. The PAST method aided collaboration with the PAB by allowing members to identify additional barriers and refine the trees based on their knowledge and experience. PASTs additionally supported the community-engaged partnership by identifying the interconnectedness of core problems (i.e., limited supply of healthy foods and preference for salty foods).

**Conclusions:** PASTs showed their relevance as tools for community-engaged research and responsive program planning. Challenges in creating and presenting intricate trees via online format highlight considerations for virtual facilitation and real-time documentation. Adaptations to the method are described for future use.

## Introduction

Problem and Solution Trees (PASTs) are relevant to community health partnerships because they facilitate shared problem identification, support collaborative decision-making, and translate community knowledge into actionable, locally responsive solutions.^1^ While PASTs have been used internationally to examine barriers related to healthy eating,^1-3^ their use in the United States (US) is limited. For example, Snowdon et al. applied PASTs across Pacific Island countries to identify nutrition policy gaps and inform the development of locally tailored interventions.^1^ They successfully identified many factors contributing to poor diets and developed feasible solutions.^1^ The solutions targeted individuals and various sectors, including banking, education, trade, and agriculture.^1^ Albert et al. used problem trees in the Solomon Islands to identify the drivers of poor dietary diversity and guide multilevel solutions focused on agriculture, education, and food systems.^2^ Similarly, Jacob Arriola et al. used PASTs in Kenya to link behavioral root causes of child stunting to practical water, sanitation, and hygiene (WASH) and nutrition strategies.^3^ Given the limited application of PASTs in the US, this paper describes how a community-academic partnership applied and adapted the PAST method to identify barriers to DASH (Dietary Approaches to Stop Hypertension) eating plan adoption and adherence among economically disadvantaged Black/African Americans (hereinafter referred to as Black Americans). This paper also provides recommendations for applying PASTs within community-engaged research and program planning.

### Community-Academic Partnership

The DASH eating plan is an evidence-based strategy for preventing and managing hypertension that promotes eating heart-healthy foods, such as fruits and vegetables, and limits sodium, saturated fat, and added sugars.^4,5^ Despite decades of evidence, the DASH plan is not

widely followed, especially by communities that bear the burden of hypertension, such as economically disadvantaged Black Americans.^6,7^ This is concerning given that hypertension affects nearly half of adults in the US, with disproportionately higher rates among Black adults.^8^

Barriers to following the DASH plan for economically disadvantaged Black Americans include the high cost of DASH-friendly foods and the limited availability of culturally relevant recipes.^7^ Overcoming these obstacles is crucial to increasing DASH adoption in these communities that are at the highest risk of hypertension. Thus, a community-academic partnership was formed to understand the barriers influencing DASH eating plan adoption among Black Americans living in a public housing community. The academic team has partnered with the local housing authority since 2018 through research and community activities with residents. This longstanding partnership provided the foundation for the current project and formation of the PAB. Building on this partnership, community residents and organizational partners were invited to join a Project Advisory Board (PAB). Potential PAB members were identified through prior research and community partnerships, including community residents, representatives from the local housing authority, and members of community-based organizations focused on food access and food equity. The academic team comprises students and researchers from the local land-grant university, including one team member who is also a community resident.

The goal of the community-academic partnership is to increase DASH adoption among economically disadvantaged Black Americans by co-creating a culturally tailored DASH cookbook featuring low-cost recipes with community members and community/organizational partners on the PAB. Given the barriers to accessing healthy foods in these communities,^9,10^ diverse perspectives on the PAB helped ensure that recipes and resources are feasible and responsive to local needs. Community and organizational partners contributed to project planning and development of the PASTs. A community organization partner also contributed to the review and development of this manuscript. Further, few studies describe participatory methods for identifying structural and contextual barriers to healthy eating in underserved communities.

Community-engaged research methods can incorporate community knowledge and lived experiences into identifying local problems and potential solutions.^11^ At the start of the project, the PAB employed PASTs to identify community-specific barriers to following the DASH plan and co-develop solutions to improve DASH adoption and adherence, including access to DASH-friendly foods, in local, predominantly Black, underserved communities. PASTs are visual tools for understanding complex environmental and health problems (e.g., WASH and malnutrition) and for identifying appropriate solutions.^1^ PASTs rely on the knowledge and experiences of community members and community partners to ensure the issues identified reflect reality and that the proposed solutions are actionable.^1^

### Applying and Adapting the Problem and Solution Tree Method

#### Ethics Review

This study was reviewed and approved by the university’s Institutional Review Board.

#### Adaptations to the PAST Method

The PAST method was adapted to align with the PAB’s time constraints, goals, and structure. The PAB applied PASTs across a series of meetings to support meaningful participation and to ensure an efficient use of time. As in previous applications of PASTs,^1-3^ several components were predefined to streamline the process. Using previous PAST examples, the research team predefined the core problem (low adherence to the DASH eating plan among economically disadvantaged Black Americans) and its immediate effect (higher hypertension rates).^7,8^ To initiate tree development, immediate causes (cost, supply, and preference) were also preidentified based on a previous PAST application examining barriers to healthy eating in the food environment and literature on barriers to DASH adherence.^1,7^ These predefined components served as starting points rather than fixed categories and could be revised, removed, or expanded by PAB members during the meetings. For example, PAB members were asked to reflect on their own experiences operating produce distribution programs in the target community and to consider how barriers to accessing those foods, as reported by residents, could be attributed to cost, supply, or preference. After group discussion, the PAB identified education and time as additional core barriers to DASH adoption that were not included among the three immediate causes initially derived from the literature. In addition, each immediate cause was developed as a stand-alone tree rather than merged into a single combined diagram, which made group discussions more focused and resulted in the creation of five trees.

#### Tools, Setting, and Participants

The academic team identified potential PAB members through prior research and volunteer service engagements with key community partners in the community of interest. Inclusion criteria for the PAB were that potential members be (1) at least 18 years of age, (2) community members who self-identified as Black and resided in a local underserved community, and/or (3) individuals who had knowledge or experience working on community-based nutrition education or working with local underserved communities. Of the 10 PAB members who enrolled, the mean age was 44.33 (standard deviation = 13.5), 90% identified as female, and 90% identified as Black. Most PAB members were affiliated with a community-based organization whose mission focused on food equity and access to fresh produce or with a local government agency (80%), while 30% were community members, with some having more than one affiliation. Each meeting included 2 to 9 PAB members and 3 to 5 research team members.

Each PAB member received a project workbook that provided background information on the project, the PAB meeting structure, and the PAST method. The workbook also included blank templates for developing PASTs (Figure 1). Meeting agendas were emailed 1-2 weeks before each meeting. PowerPoint was used to guide the discussion, and the trees were developed in real-time on shared slides during the meeting. Meetings were conducted in person at a convenient community location or via Zoom, based on member preferences, and lasted approximately 90 minutes.

**Figure 1:**
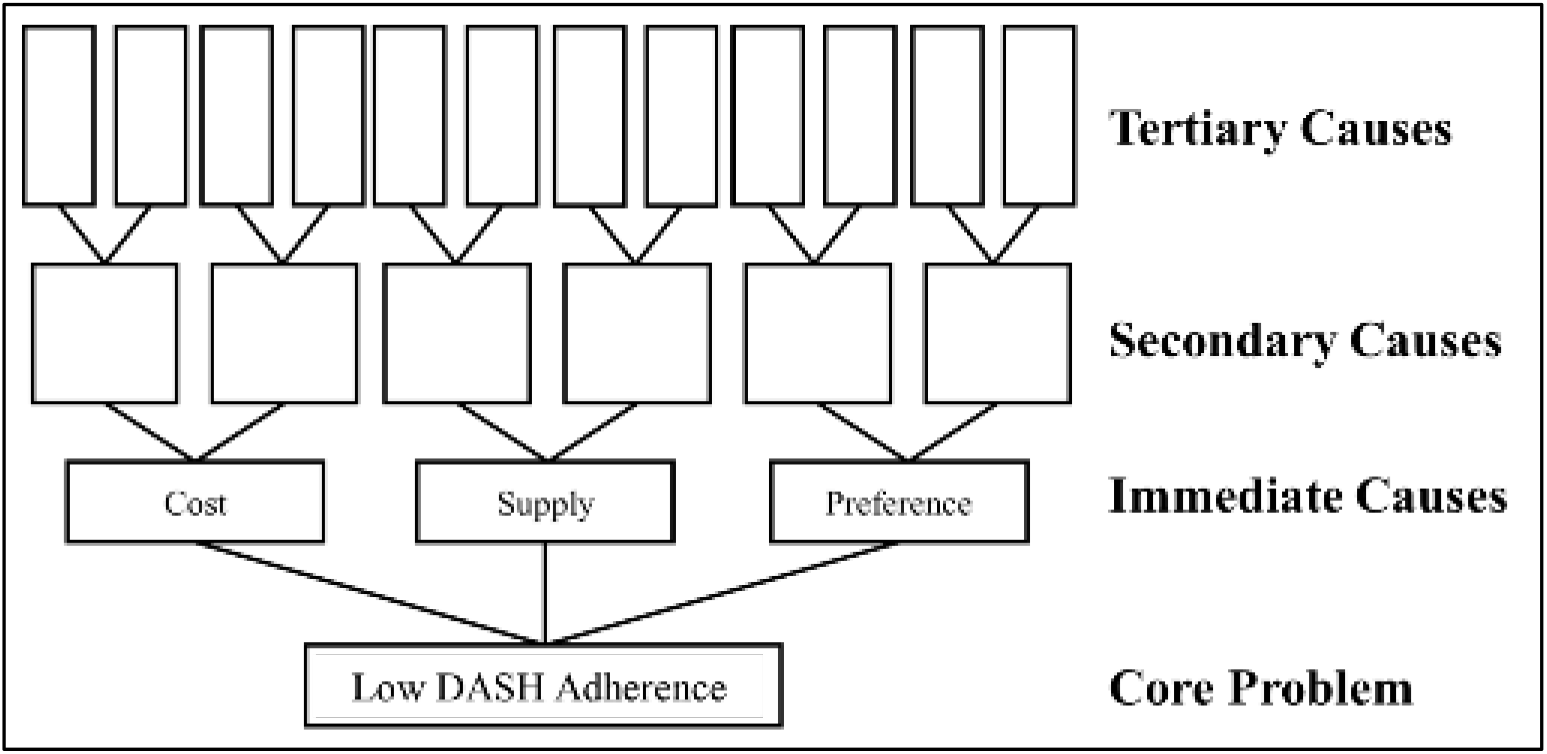
Blank template for problem tree construction provided in the PAB workbook

#### PAST Meeting Procedures

PAST activities were completed across five meetings over seven months (Table 1).

**Table 1:** Meeting activities for the construction of problem and solution trees (PAST)

| **Meeting** | **Activities** |
|---|---|
| 1 | Introduction to the PAST method and Project Advisory Board (PAB) workbook (virtual) |
| 2 | Construction of problem trees for each immediate cause (cost, supply, preference, education, and time); group discussion (virtual) |
| 3 | Construction of solution trees for cost and supply; group discussion and consensus (virtual) |
| 4 | Construction of solution trees for preference, time, and education; group discussion and consensus (virtual) |
| 5 | Review, refinement, and final approval of problem and solution trees (in-person) |

Adjustments and edits were made where needed. Meeting 1 introduced the PAB workbook and outlined the PAST method. In Meeting 2, members used the preidentified core problem, immediate effect, and immediate causes (or barriers) to construct individual problem trees for each immediate cause. During this meeting, the PAB added two additional immediate causes: education and time. For each immediate cause, PAB members identified secondary and tertiary causes. Questions such as “What is the cause of the immediate cause?” and “What factors contribute to this problem?” were used to facilitate group discussions. The academic team members began the meetings by reviewing the tree created in the previous meeting, then invited PAB members to add commentary, if necessary, to further enhance the tree or move forward with the next tree. Rather than a strict question-and-answer format, meetings were characterized by organic conversation between the academic team and PAB members, with the academic team also providing input that contributed to the construction of the tree. Following each meeting, the academic team reviewed audio transcripts to ensure the accuracy of the trees created in real time.

Solution trees were constructed in Meetings 3 and 4 by identifying one or more potential actionable solutions for each secondary or tertiary cause (Figure 2). Meeting 3 focused on cost and supply, while Meeting 4 addressed preference, time, and education. In Meeting 5, the PAB reviewed and approved the complete set of trees and agreed that they would remain living documents for future planning and action.

**Figure 2:**
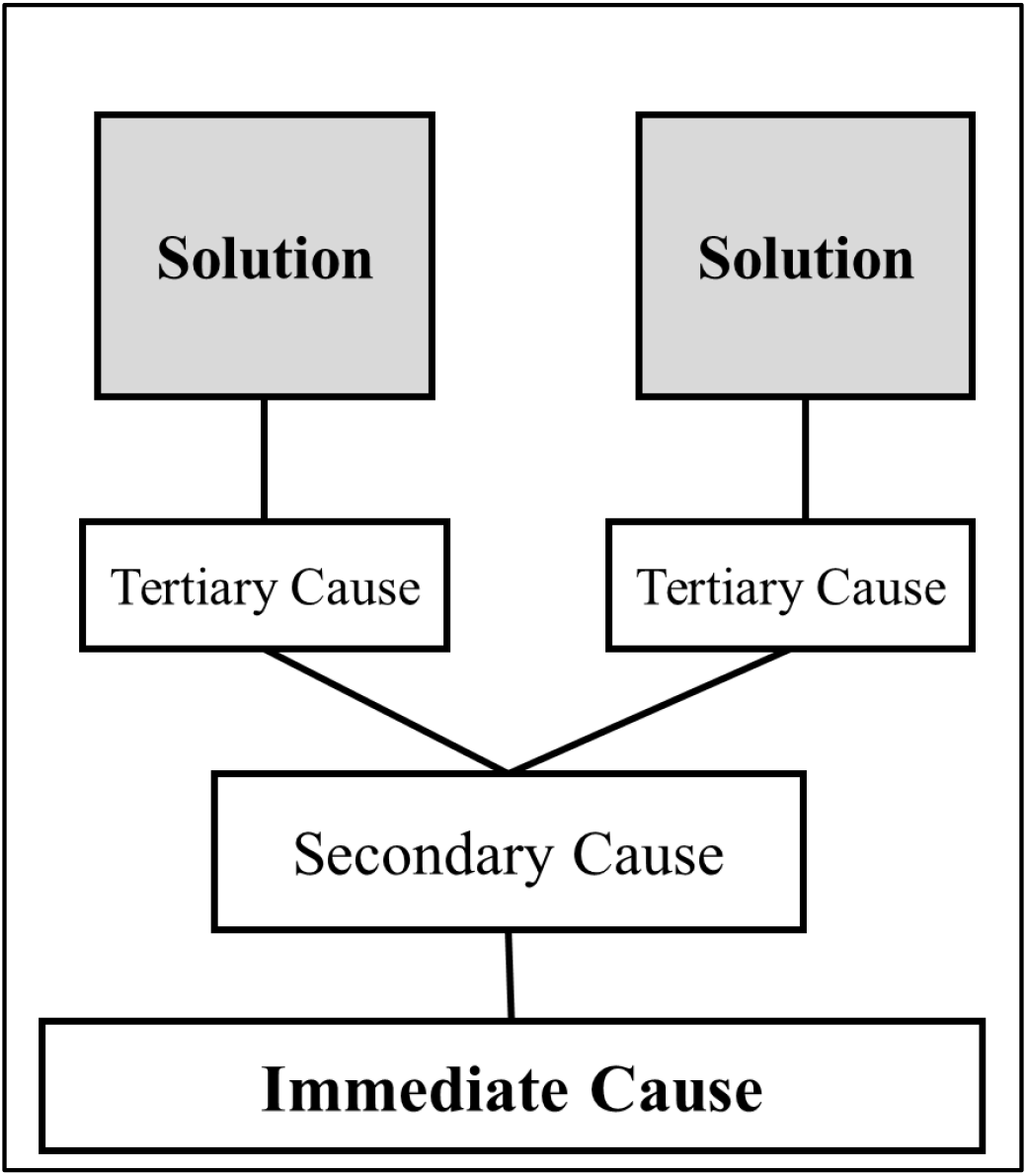
Template for solution trees constructed in meetings 3-4

#### Data Documentation and Consensus

During meetings, the research team documented PAB members’ input on the PAST diagrams in real time on a shared PowerPoint screen. Meeting minutes were taken by project research assistants and distributed to PAB members after each meeting. All meetings were recorded, transcribed, and deidentified. Transcripts and meeting minutes/notes were reviewed to ensure the trees captured all PAB input. The transcripts and meeting documentation were used to verify the content of the trees. Updated trees were revisited during subsequent PAB meetings to reach group consensus. Consensus was achieved through group discussion, during which PAB members reviewed each tree, proposed edits as needed, and confirmed agreement before finalizing the trees. Meeting minutes were shared with the PAB two weeks following each meeting. Thus, members who missed the meeting would be able to review the discussion highlights and provide feedback at the following meeting.

### Lessons Learned and Considerations for Future Use

Applying PASTs with our PAB identified several considerations related to facilitation, community participation, managing complex trees, and use of the method for program planning.

#### Facilitation and Documentation

The visual format of the trees helped participants identify overlapping issues and duplicative efforts, and helped the PAB identify actionable solutions. However, displaying and editing trees in real time via shared PowerPoint slides over Zoom was sometimes difficult, particularly when trees included numerous secondary causes, making them visually complex and hard to interpret on screen (Figure 3). Creating complex trees in real time was challenging, especially with only one to two members of the research team available to make edits. For future applications, using collaborative virtual tools such as Lucidchart or Slido may make collaboration easier when creating the trees. In addition, conducting tree development meetings in person may enable more collaborative editing, helping identify further secondary and tertiary causes and solutions.

**Figure 3:**
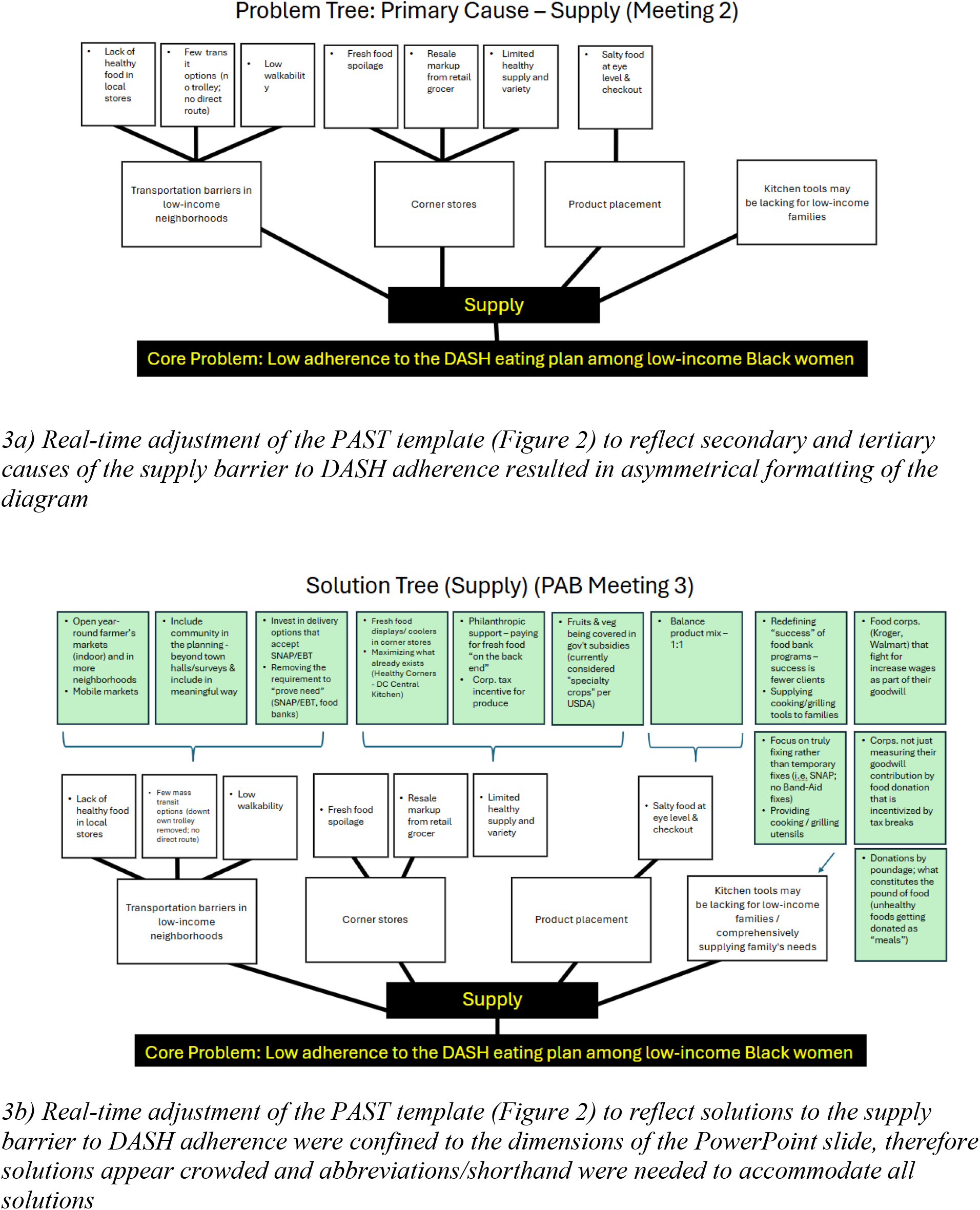
PAST for supply created in real-time during PAB meetings 2 (problem) and 3 (solution)

However, it is important to consider participants’ preferences and comfort with technology, as well as their availability to meet in person over a series of meetings to construct the trees. Future academic teams contemplating the use of PASTs should also maintain flexibility in the number of meetings with the community, whether virtual or in-person, that may be required to achieve complete trees. For instance, the initial literature focused on three core barriers to DASH adherence, while the PAB identified education and time as additional barriers, requiring five meetings rather than three.

#### Balancing Structure and Community Input

Predefining the core problem, immediate effect, and immediate causes from prior PAST applications and DASH literature reduced the time and effort needed to examine the complexity of structural inequities in accessing healthy foods. This approach allowed the PAB to make early progress on tree construction and created an initial sense of accomplishment. At the same time, PAB members had the opportunity to modify and expand the predefined components, thereby fostering shared ownership.

Revisiting and refining the trees across multiple meetings improved their accuracy and quality. This iterative process also promoted shared ownership by allowing participants to reconsider earlier ideas, suggest new ideas, and clarify relationships between causes and solutions. This was especially important because several of the trees were complex and multilevel.

#### Managing Complexity Across Trees

Constructing separate trees for each immediate cause (e.g., cost, supply, time, education, and preference) had both advantages and drawbacks. Separating the trees allowed the PAB to explore each barrier in depth and maintain focused discussions. However, as additional trees were constructed, overlap emerged across causes, highlighting the interconnectedness of factors contributing to low DASH adherence among Black Americans. For future use, PAST facilitators should plan time for cross-tree synthesis to identify shared root causes and solutions.

#### Supporting Community-Engaged Planning

The PAST method provided a process for identifying local community needs. PAB members’ shared interests in improving cardiovascular health among economically disadvantaged Black families helped build agreement around key barriers and priorities. Working collaboratively with PAB members to construct the trees helped identify unrecognized problems, prioritize community needs, and identify duplicative efforts. The process also helped identify opportunities for PAB member organizations to build new partnerships and/or enhance ongoing initiatives. For example, one PAB member who operated a mobile produce distribution program expanded their schedule to distribute produce to the public housing community managed by another PAB member. PAST discussions were also incorporated into FoodChain’s grocery store planning, including considerations of store layout, design, and community engagement. These discussions helped translate identified barriers and potential solutions into practical decisions about how the space and its services could better respond to community needs. The PAST method functioned as both a research and community program planning tool. The completed trees have informed project activities, generated new program ideas, and shaped future research directions by linking identified barriers with potential solutions and relevant community or organizational partners.

For example, one problem identified by the PAB was a lack of kitchen resources and supplies needed to follow the DASH eating plan. The researchers subsequently received funding for an implementation science research study to co-create a kitchen toolkit with the community to support DASH adoption. This demonstrates how PASTs can support both problem identification and subsequent community-engaged planning.

## Conclusions

The PASTs provided a participatory, visual approach to identifying community-specific barriers to DASH adoption and to collaboratively developing potential solutions. Specifically, our team applied PASTs with community members and organizational partners in the US. The PAST method enabled our team, in collaboration with the PAB, to identify community needs, local determinants of health, and pathways that contribute to health inequities. The process supported shared problem identification and decision-making by allowing PAB members to modify the trees, identify additional barriers, and collaboratively develop potential solutions. Our experience also demonstrated the importance of allowing sufficient time and flexibility for community input and adapting facilitation approaches as tree development becomes more complex. Future applications should also consider how connections across multiple trees can be examined to identify common barriers and opportunities for action.

## Acknowledgements

We are grateful to the Project Advisory Board (PAB) members for their time, insights, and ongoing partnership throughout this work. We especially thank KH, a PAB member, for her contribution to the manuscript review and for her input. Research reported in this publication was supported by the National Library of Medicine of the National Institutes of Health under Award Number G08LM014412. The content is solely the responsibility of the authors and does not necessarily represent the official views of the National Institutes of Health.

## Data Availability Statement

The data supporting the findings of this study are not publicly available due to privacy and confidentiality considerations associated with the small sample of community stakeholders.

